# Performance of Fabrics for Home-Made Masks Against the Spread of Respiratory Infections Through Droplets: A Quantitative Mechanistic Study

**DOI:** 10.1101/2020.04.19.20071779

**Authors:** Onur Aydin, Bashar Emon, Shyuan Cheng, Liu Hong, Leonardo P. Chamorro, M. Taher A. Saif

## Abstract

Respiratory infections may spread through droplets and aerosols released by infected individuals coughing, sneezing, or speaking. In the case of Coronavirus Disease 2019 (COVID-19), spread can occur from symptomatic, pre-symptomatic, and asymptomatic persons. Given the limited supply of professional face masks and respirators, the U.S. Centers for Disease Control and Prevention (CDC) has recommended home-made cloth face coverings for use by the general public in areas of significant community-based transmission. There is, however, little information on the effectiveness of cloth face coverings in reducing droplet dissemination. Here, we ascertained the performance of 11 household fabrics at blocking high-velocity droplets, using a commercial medical mask as a benchmark. We also assessed their breathability (air permeability), texture, fiber composition, and water absorption properties. We found that droplet blocking efficiency anti-correlates with breathability; less breathable fabrics being more effective in blocking. However, materials with high breathability are desirable for comfort and to reduce airflow through gaps between the mask and face. Our measurements indicate that 2 or 3 layers of highly permeable fabric, such as T-shirt cloth, may block droplets with an efficacy similar to that of medical masks, while still maintaining comparable breathability. Overall, our study suggests that cloth face coverings, especially with multiple layers, may help reduce droplet transmission of respiratory infections. Furthermore, face coverings made from biodegradable fabrics such as cotton allow washing and reusing, and can help reduce the adverse environmental effects of widespread use of commercial disposable and non-biodegradable facemasks.

## Introduction

At the crux of any pandemic is the transmission of a pathogenic agent, *e.g*., a novel virus that spreads in populations on a global scale. Current knowledge from influenza, SARS-1, and MERS indicates three major routes of transmission, namely, droplets, aerosols, and contact.^1,2^ Although the mechanism of spread of the current novel coronavirus (SARS-CoV-2) is not clearly understood, it is thought that spread can occur through droplets containing virus particles released by infected persons when they sneeze, cough, or speak.^3,4^ Larger droplets tend to fall nearby by gravity, and the sufficiently smaller ones can stay in the air and travel longer.^1,5,6^ Droplets inhaled by a healthy individual allow the virus to enter the respiratory system and cause infection. Face masks can offer a physical barrier against virus transmission. This is particularly true for SARS-CoV-2 viruses that are shed by symptomatic, pre-symptomatic, and asymptomatic carriers.^7–10^

During the COVID-19 pandemic, the supply of commercially manufactured facemasks has not been able to meet the demand. The U.S. CDC has therefore provided guidance for the public to use non-commercial alternatives to facemasks such as cloth face coverings to slow the spread of COVID-19.^11^ However, it is not yet clear what kind of fabric would be the most efficient material or how many layers of cloth would protect against both spreading and contracting the virus. Existing literature on home-made masks have mostly focused on the filtration efficacy of the masks against aerosol or dry airborne particles less than 10µm in size under prescribed flows or pressure differentials.^12–16^ These studies are relevant for situations where an individual is breathing in an atmosphere with pathogenic aerosol particles. In contrast, when an infected individual coughs, sneezes, or talks into a mask, the droplets that would hit the inside of the mask are relatively large, and have high momentum.^6^ It is also possible (especially for healthcare workers and family members in contact with infected individuals) to be on the receiving end of such droplets that may be loaded with pathogens. How home-made masks can be effective in these scenarios remains elusive. To address this issue in the current context of COVID-19, we evaluated 11 regular household fabrics with varying texture and fiber compositions, using a commercial medical mask as a benchmark. We measured their efficiency at blocking high-velocity droplets and their breathability (*i.e*., air permeability) in a laboratory setting. To provide a mechanistic understanding of how household fabrics may block droplets, we also assessed their texture, fabric type, and wetting, and water absorption behavior.

In this study, we did not consider steps for producing a facemask, *e.g*., how they should be stitched, how their boundaries should be designed, how to attach them to the face, and how they should be used or decontaminated. For detailed information on how to make, use, and decontaminate cloth face coverings, we refer the reader to guidelines provided by the U.S. CDC and World Health Organization (WHO).^11,17–19^ Additionally, filtration efficiency of various fabrics against aerosolized and dry airborne particles (usually less than 10 µm in size) has been studied previously,^15,16,20^ and is beyond the scope of this study. Our goal here is to quantify the effectiveness of household fabrics at blocking larger droplets and offer insight into the possible mechanisms of droplet blocking.

## Results and Discussion

### Problem Definition

To understand the effectiveness of any mask against high-velocity droplets carrying 100 nm particles, we asked the following questions: (1) what are the essential parameters in determining the mask material against droplets? (2) what is the physical mechanism by which a mask material can block the droplets? To answer the first question, we note that there are two groups of mask users: (a) infected individuals releasing droplets by sneezing, coughing, and speaking, and (b) healthy individuals receiving the droplets. For the former, the mask is most challenged when droplets land on the mask with high momentum (high mass and velocity).^21,22^ For the recipient, the droplet momentum is small or negligible. If a common mask is to be applicable to both, then the mask material must be able to resist both high and low-momentum droplets. The mask should also be breathable (*i.e*., have sufficient air permeability) so that air does not go through the gaps between the mask and face. A large fraction of airflow through the gaps defeats the purpose of the mask, as droplets can be carried with the side flow, and the mask provides a false sense of protection. Thus, we consider the droplet blocking efficiency, *ε*, and the breathability, *β*, as the two critical parameters for mask materials. It is important to note that while high breathability is desirable, higher air permeability may correspond to lower droplet blocking efficiency. The problem of finding an appropriate material for a home-made mask thus involves choosing a fabric that has high droplet blocking efficiency while still offering sufficient breathability.

Here, we considered a diverse set of 11 common household fabrics, and used a medical mask as our benchmark (Figure 1). The household fabrics were selected from new and used garments, quilt cloths, bed sheet, and dishcloth, and characterized in terms of their fabric construction (woven, knit, or napped), fiber content (cotton, polyester, polyamide, silk), weight, thread count, and porosity (see Methods). Sample descriptions and parameter values are listed in Table 1.

**Table 1.** Characterization of fabrics used in the study.

| **Sample Description** | **Fabric Construction** | **Fiber Content** | **Weight (g/m<sup>2</sup>)** | **Thread Count (threads per inch)** | **% Porosity, mean ±SD (n=9)** |
| --- | --- | --- | --- | --- | --- |
| Medical Mask: FM-EL style | non-woven | polypropylene | 53.9 | n/a | n/a |
| Fabric 1: Used shirt | knit | 100% cotton | 114.2 | 200 | 0.7 ±0.1 |
| Fabric 2: New undershirt | knit | 100% cotton | 111.5 | 45 | 4.5 ±0.1 |
| Fabric 3: New quilt cloth | woven | 100% cotton | 89.1 | 150 | 10.8 ±0.2 |
| Fabric 4: Used undershirt | knit | 75% cotton, 25% polyester | 148.2 | 85 | 5.5 ±0.2 |
| Fabric 5: Used shirt | woven | 70% cotton, 30% polyester | 107.5 | 200 | 0.1 ±0.1 |
| Fabric 6: New T-shirt | knit | 60% cotton, 40% polyester | 183.2 | 75 | 1.1 ±0.3 |
| Fabric 7: New quilt cloth | woven | 35% cotton, 65% polyester | 95.4 | 180 | 4.8 ±0.2 |
| Fabric 8: new bedsheet | woven | 100% polyester | 81.1 | 180 | 5.8 ±0.8 |
| Fabric 9: New dishcloth | napped | 80% polyester, 20% polyamide | 380.5 | n/a | n/a |
| Fabric 10: Used shirt | woven | silk | 49.9 | 220 | 4.3 ±0.1 |
| Fabric 11: Used shirt | woven | silk | 49.4 | 200 | 2.2 ±0.5 |

**Figure 1.**
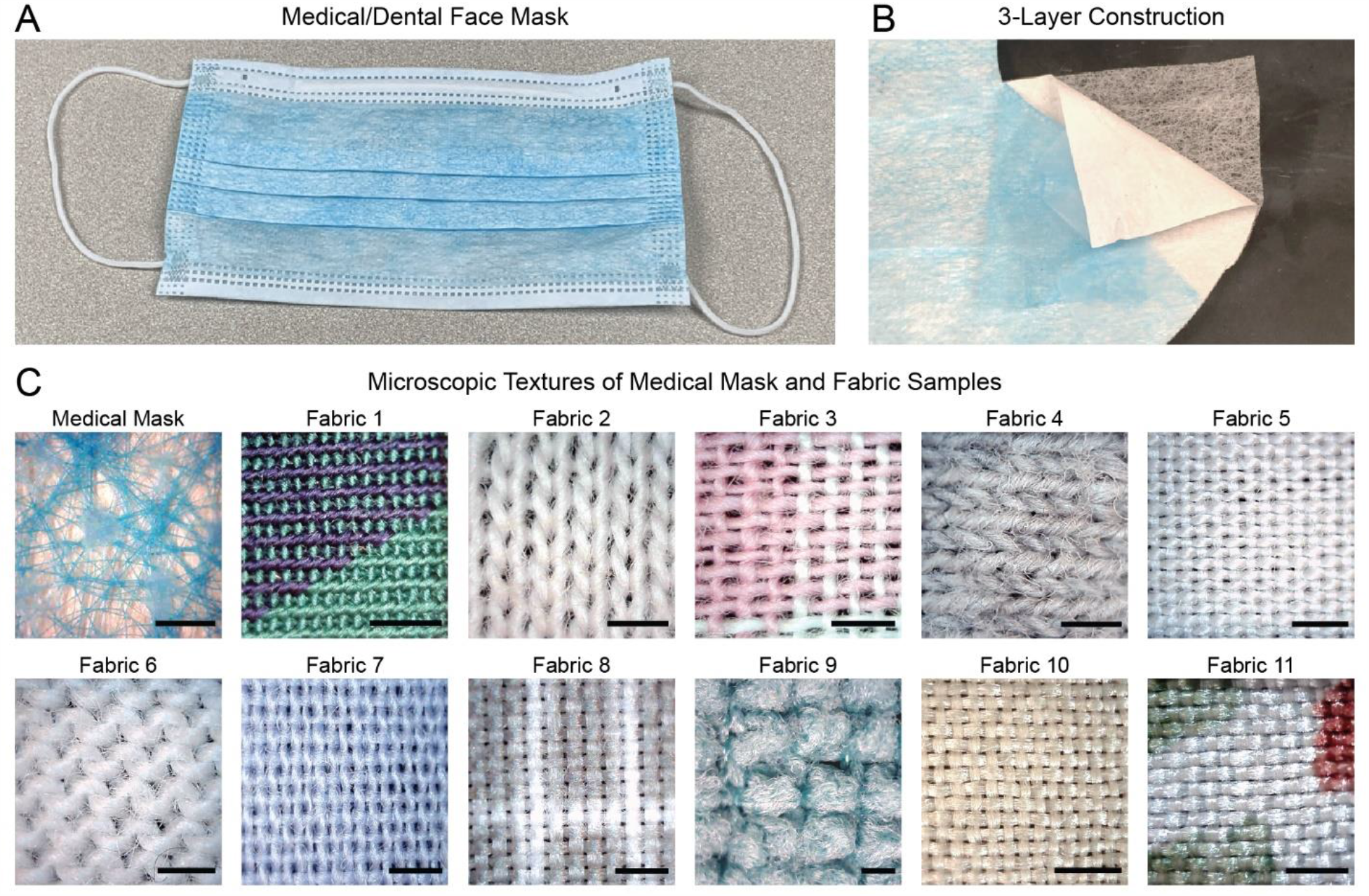
Samples used in this study. (A) Image of a medical/dental quality FM-EL style face mask with (B) 3-layer construction, which was used as a benchmark. (C) Microscopic texture of the outer surface of the medical mask and the 11 different home fabric samples. All scale bars: 1 mm.

### Droplet Blocking Efficiency

To ascertain the possible utility of the chosen fabrics as materials for cloth face coverings, we first developed a method to challenge the fabrics with high and low-velocity droplets and to quantify their droplet blocking efficiency. A schematic of our experimental method is shown in Figure 2A. To generate droplets with a high initial velocity, we repurposed a metered-dose inhaler. Such inhalers, when pressed, generate sprays with consistent ejection pressure and duration.^23–25^ We loaded the nozzle of the inhaler with a suspension of 100 nm-diameter fluorescent beads in distilled water. The fluorescent beads serve two purposes: (1) mimic SARS-CoV-2 virus (70-100 nm-diameter)^26,27^ in terms of size, and (2) allow to quantify the blocking efficiency of the fabric samples. When the inhaler is pressed, the internal pressure of the inhaler ejects bead suspension out of the nozzle, creating high-speed droplets (Video S1). The droplets then hit the fabric sample that is placed in front of the inhaler (Figure 2B).

**Figure 2.**
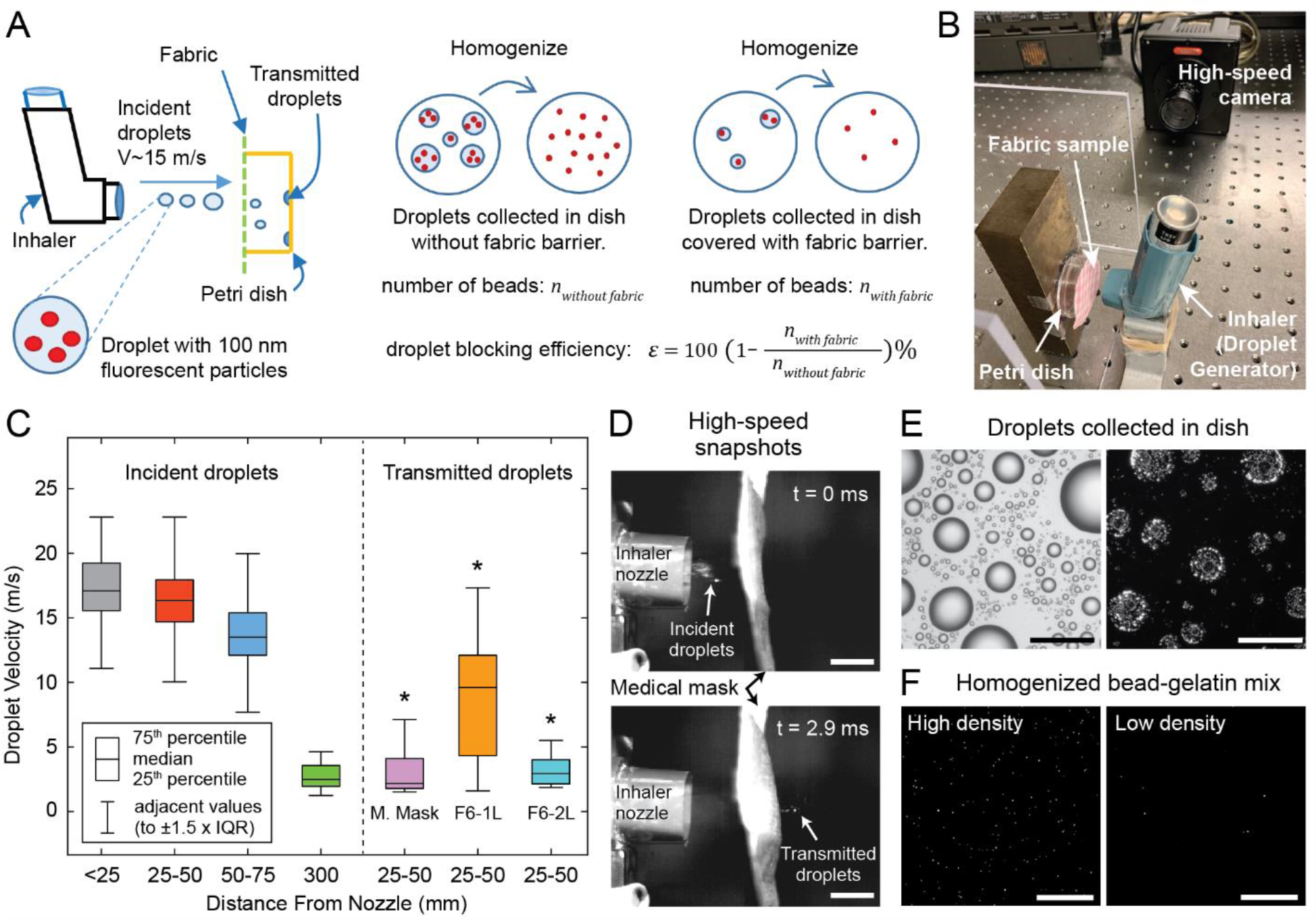
Droplet challenge tests. (A) Schematic of the experimental method and (B) image of lab set-up. (C) Box plots showing incident droplet velocities at various distances from the inhaler nozzle, and exit velocities of droplets that penetrate medical mask and single and double-layered T-shirt fabric (fabric 6). *p<0.001, two-sample t-test comparing exit velocities of penetrating droplets to velocities of free droplets in 25-50 mm range. (D) High-speed snapshots of droplets hitting and penetrating the medical mask. Scale bars: 10 mm. (E) Brightfield and fluorescence images of droplets collected on a petri dish. Scale bars: 100 µm. (F) Confocal images of homogenized bead collection; representative images from samples with high and low bead density. Scale bars: 100 µm.

We recorded high-speed videos of the ejected droplets and performed image analysis to estimate droplet size and velocity (see Methods for details). We detected droplets with diameters in the ∼0.1 mm to ∼1 mm range within the spray ejected by the inhaler (Figure S1). Droplets exit the inhaler with a median velocity of 17.1 m/s, measured within 25 mm (∼1 inch) from the nozzle, and gradually slow down as they travel through the air. Median droplet velocity reduces to 2.7 m/s after they travel 300 mm (∼12 inch) from the inhaler nozzle (Figure 2C). Droplets released by sneezing, coughing, and speaking typically vary in the ranges of 0.5 µm to 1 mm in size and 6 m/s to 100 m/s in velocity.^6,28–31^ Hence, our experimental platform falls within the range of these respiratory events, and, in contrast to aerosol filtration studies, offers a way to test blocking efficiency against larger (up to ∼1 mm size) droplets. We chose 25 mm from the inhaler nozzle as the location to place the fabric samples to challenge them with high-momentum droplets. High-speed video recordings verify that droplets can penetrate the medical mask and fabrics with 1 or 2 layers at this range (Figure 2D, Videos S2-S4). We also quantified the velocities of the droplets that penetrate the medical mask and 1 and 2 layers of T-shirt fabric (fabric 6 in Table 1) and compared them to the velocities of free droplets within the same region (25 to 50 mm from the inhaler nozzle). Droplets penetrating a single layer of T-shirt fabric had a median velocity of 9.6 m/s. In contrast, droplets penetrating a medical mask and 2 layers of T-shirt fabric had median velocities of 2.2 m/s and 3.0 m/s, respectively (Figure 2C). In all three of these cases, velocities of the droplets that penetrated the fabric barriers were significantly lower than the incident velocity of 17.1 m/s (p < 0.001 for all, two-sample t-test).

To collect the droplets that were transmitted through the fabrics, we placed a petri dish behind the fabric samples (Figure 2A,B). Similarly, petri dish without a fabric barrier was used at the same location to collect incident droplets for comparison. Figure 2E shows brightfield and fluorescence images of droplets that were collected in the petri dish without a fabric barrier. Since fluorescent beads are uniformly dispersed in the solution that is loaded into the inhaler nozzle, we can use the number of fluorescent beads collected with and without a fabric barrier to measure the droplet blocking efficiency. However, the droplet images in Figure 2E cannot be used to count the beads because many beads are clustered together and cannot be counted separately. To solve this problem, we developed a method to disperse and homogenize the beads collected in the petri dish. We achieved homogenization by first collecting the droplets on petri dishes coated with a layer of gelatin hydrogel, then melting the gelatin at 37°C, mixing the beads with liquid gelatin, and re-gelling. We used a confocal microscope to image the gelatin hydrogels containing fluorescent beads (Figure 2F). For each test, 100 to 150 images were taken from separate locations of the hydrogel and the average number of beads per image, 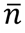, was computed by image analysis. We carried out control measurements to validate our method and found that 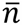 can be used as an accurate predictor of the bead density in the gelatin mixture within the range of 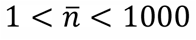 (Figure S2, see Methods for details of bead mixture homogenization, confocal imaging, analysis, and validation).

To measure droplet blocking efficiency, we need to compute a ratio of the total number of beads transmitted through the fabric and collected in the gelatin-coated dish, *n*_*with fabric*_, to the total number of beads collected without a fabric barrier, *n*_*without fabric*_. To compute this ratio, we also took into account the total volume of gelatin solution, *V*, that the fluorescent beads were dispersed in for each test. Since the average number of beads per image, 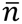, correlates with bead density (*i.e*., number of beads per unit volume), we use 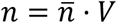 as a predictor of the total number of beads collected in the petri dish. Droplet blocking efficiency, *ε*, of the fabric is thus computed as:

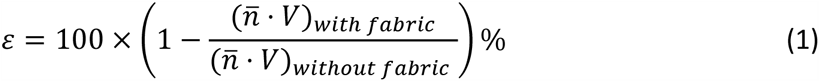

We performed independent measurements of 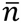 without any fabric barrier (15 separate measurements, 100-150 images each), as well as with the medical mask, all 11 household fabrics with a single layer, and fabric 2 and fabric 6 (see Table 1) with 2 and 3 layers (6 or 7 separate sample measurements for each fabric, 100-150 images per measurement). We then computed the droplet blocking efficiency, *ε*, using Equation 1, for all possible combinations of 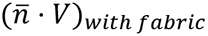 and 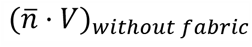. Distributions of droplet blocking efficiency values for all tests are shown in Figure 3. The minimum, median, and maximum values are provided in Table 2.

**Table 2.** Results of droplet blocking efficiency (*ε*) and breathability (*β*) measurements.

| Sample description | $\varepsilon$ (%) at 25 mm (high momentum droplets) | | | $\beta$ (mm/Pa·s) |
| --- | --- | --- | --- | --- |
| | Minimum* | Median | Maximum | Mean ±SD |
| **Medical Mask** | 96.4 | 98.5 | 99.9 | 1.83 ±0.15 |
| **Fabric 1:** Used shirt, knit, 100% C | 87.9 | 96.8 | 99.8 | 1.37 ±0.06 |
| **Fabric 2:** New undershirt, knit, 100% C | 41.1 | 81.9 | 95.2 | 10.70 ±0.66 |
| **Fabric 3:** New quilt cloth, woven, 100% C | 30.6 | 71.7 | 93.3 | 8.67 ±0.12 |
| **Fabric 4:** Used undershirt, knit, 75% C - 25% PE | 28.9 | 72.5 | 92.5 | 11.97 ±0.25 |
| **Fabric 5:** Used shirt, woven, 70% C - 30% PE | 81.2 | 93.6 | 99.7 | 1.80 ±0.00 |
| **Fabric 6:** New T-shirt, knit, 60% C - 40% PE | 42.0 | 83.1 | 98.3 | 7.23 ±0.55 |
| **Fabric 7:** New quilt cloth, woven, 35% C - 65% PE | 55.2 | 81.8 | 96.4 | 5.07 ±0.21 |
| **Fabric 8:** New bedsheet, woven, 100% PE | 74.9 | 94.8 | 99.7 | 3.23 ±0.06 |
| **Fabric 9:** New dishcloth, napped, 80% PE - 20% PA | 90.0 | 98.2 | 99.8 | 6.53 ±0.21 |
| **Fabric 10:** Used shirt, woven, 100% S | 70.8 | 92.9 | 99.5 | 3.90 ±0.36 |
| **Fabric 11:** Used shirt, woven, 100% S | 81.1 | 98.7 | 99.8 | 2.10 ±0.61 |
| **Fabric 2 - 2 Layers** | 78.3 | 94.1 | 98.3 | 5.53 ±0.35 |
| **Fabric 2 - 3 Layers** | 96.8 | 98.9 | 99.8 | 3.77 ±0.06 |
| **Fabric 6 - 2 Layers** | 94.0 | 98.1 | 99.6 | 3.87 ±0.06 |
| **Fabric 6 - 3 Layers\*\*** | | >98.1 | | 2.63 ±0.06 |
| Sample description | $\varepsilon$ (%) at 300 mm (low momentum droplets) | | | |
|  | Minimum* | Median | Maximum |  |
| **Medical Mask** | 95.2 | 99.7 | 99.9 |  |
| **Fabric 6** | 82.5 | 94.2 | 99.3 |  |
| **Fabric 6 - 2 Layers\*\*** |  | >94.2 |  |  |

C: cotton, PE: polyester, PA: polyamide, S: silk
\*Outliers (see Figure 3) are ignored. Minimum values tabulated correspond to the lower ends of whiskers in the box plots of Figure 3. (Maximum values are data maxima. There were no outliers in the upper quartiles.)
\*\*For these tests, the average number of fluorescent beads per image was below the detection limit. Hence, the expected value based on the median efficiency of one fewer layer of the same fabric is reported.

**Figure 3.**
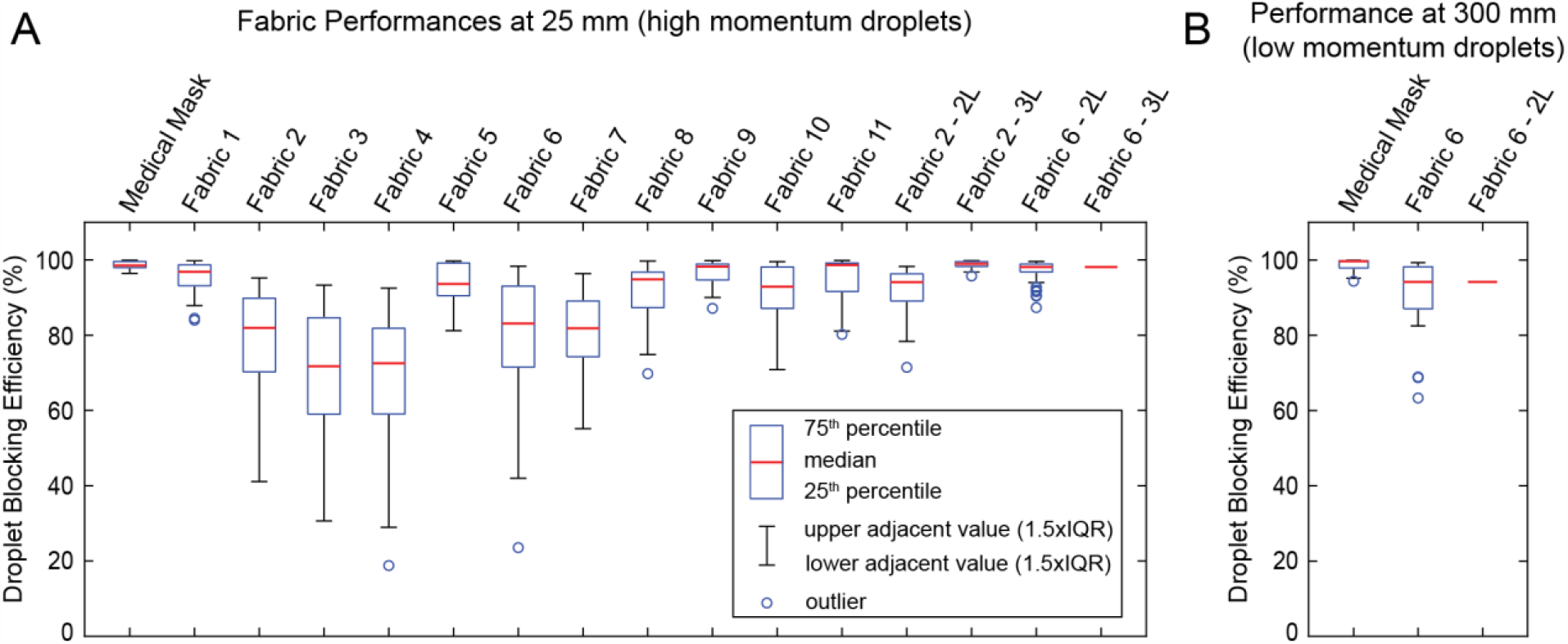
Droplet blocking efficiency. Box plots showing the distribution of droplet blocking efficiencies for different fabrics at (A) 25 mm and (B) 300 mm from the nozzle, representing efficiencies against high velocity and low velocity droplets, respectively. For 3 layers of fabric 6 at 25 mm and 2 layers of fabric 6 at 300 mm, measurements were below detection limit, therefore, only a line corresponding to the estimated lower bound is presented for these samples.

The tests at 25 mm from the inhaler nozzle represent the case of mask users releasing droplets with high-velocity onto the mask by sneezing, coughing, or speaking. At this range, we found that most fabrics with a single layer have relatively high droplet blocking efficiencies (median values > 70%) (Figure 3A, Table 2). The median droplet blocking efficiency was 98.5% (minimum 96.4%) for the medical mask. For single layers of fabrics 2 and 6, median droplet blocking efficiency was 81.9% (minimum 41.1%) and 83.1% (minimum 42.0%), respectively. The addition of second and third layers increased the efficiency, with median values exceeding 94% (Table 2). Note, however, that for 3 layers of fabric 6, the average number of beads per image was below the detection limit 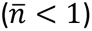. We assume that the droplet blocking efficiency of 3 layers of fabric will be at least as high as the efficiency of 2 layers (median 98.1% for fabric 6), and hence report the median efficiency for 3 layers of fabric 6 as >98.1% (Table 2). For all other tests at 25 mm,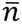 was within the accurate detection range 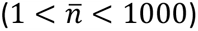.

We expected that droplet blocking efficiency would be higher for low-momentum droplets, which represents the case for users receiving droplets from a long distance. To test this expectation, we measured *ε* for the medical mask and 1 and 2 layers of fabric 6 at a distance of 300 mm from the inhaler nozzle. At this distance, high-speed imaging of the droplets shows a median velocity of 2.7 m/s (Figure 2C, Video S5). The droplet sizes at this distance are also smaller (Figure S1). Thus, the impact of the droplets has decreased. At 300 mm, median values of *ε* for the medical mask and single layer of fabric 6 were 99.7% and 94.2%, respectively (Figure 3B, Table 2). As expected, *ε* at 300 mm is higher compared to that at 25mm. For 2 layers of fabric 6 at 300mm, the average number of beads per image was below the detection limit 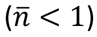; Hence, as above, we report the median efficiency as >94.2% (*i.e*., efficiency of 2 layers at least as high as the efficiency of a single layer).

We would like to stress that the efficiency values we report here are indicative of the ability of the fabric materials to block large droplets, and are not meant to reflect the full performance of a cloth facemask. The performance of a facemask would also depend on how it is worn, and whether it is also challenged by aerosols or small airborne particles (∼10 µm or smaller in size). As mentioned earlier, these questions are outside the scope of this study.

### Fabric Breathability

We define breathability, *β*, of a fabric as, *β*=*df*/*dp*, where *df* is a change in the flow rate of air through unit area of fabric, and *dp* is the corresponding change in the pressure differential across the sample that is required to induce *df*. We used a plug flow tube ^32,33^ to measure *β*. Here, the sample fabric seals the opening of a plug flow tube (Figure 4A,B). Pressurized air was pumped through the tube. Pressure outside the tube is atmospheric and the pressure inside the tube was measured with respect to the atmosphere. Thus, air was forced through the fabric by the gauge pressure, *p*. Since the area of the fabric sample subjected to air flow is the same as the cross-sectional area of the plug flow tube, the change in flow rate through unit area of fabric, *df*, is equivalent to the change in average flow velocity, *dv* inside the tube. Hence, breathability can be written as *β*=*dv*/*dp*. In a short plug tube, flow velocity is approximately uniform across the tube cross section ^33,34^. We measured velocity, *v*, at the center of the tube, while the gauge pressure, *p*, was measured at around 1/3 radius from the center. Note that the velocity profile remains the same along the length of the plug tube, while pressure is uniform at any cross section orthogonal to the tube, but it decreases along the tube, giving a pressure gradient.

**Figure 4.**
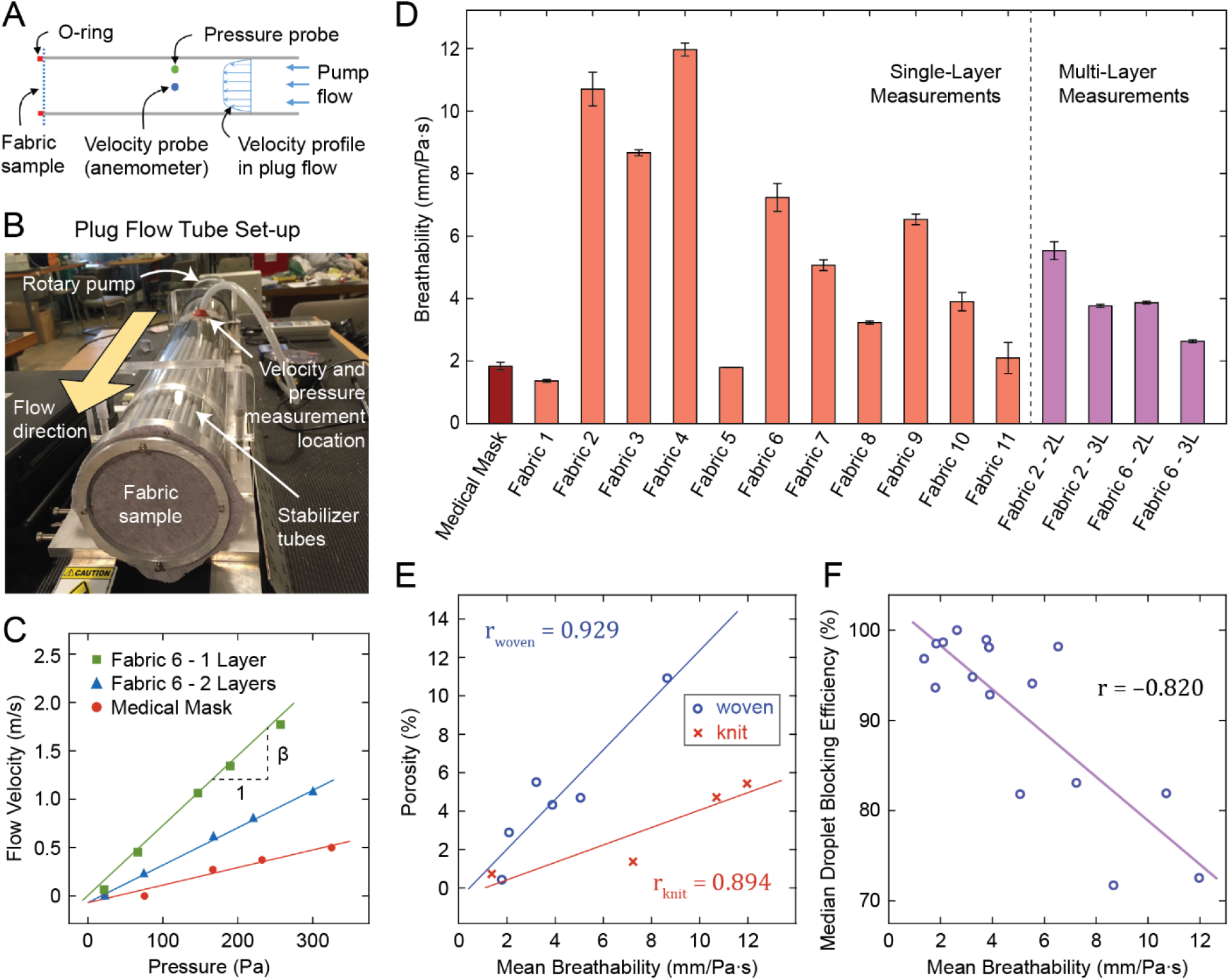
Fabric breathability measurement. (A) Schematic of the experimental method and (B) image of plug flow tube set-up. (C) Flow velocity vs. pressure plots for selected samples. (D) Breathability measurement results for all samples. (E) Fabric porosity vs. breathability for woven and knit fabrics, and (F) droplet blocking efficiency vs. breathability plots with regression lines.

Figure 4C shows velocity vs. pressure measured at various pump speeds for a single and double layered T-shirt (fabric 6) and the medical mask. This measurement was taken for the medical mask, single layers of all 11 household fabrics, as well as 2 and 3 layers of fabric 2 and fabric 6 (3 independent samples tested for each case). In all cases, velocity vs. pressure showed a linear relationship. We computed breathability, *β*, as the slope of the velocity vs. pressure plots. Results are shown Figure 4D and Table 2.

We expect that the breathability (air permeability) of fabrics would depend strongly on their porosity. Furthermore, physical intuition suggests that breathability and droplet blocking efficiency may be anti-correlated, *i.e*., the higher the breathability the lower the blocking efficiency. To assess these relationships, we plotted fabric porosity and droplet blocking efficiency against breathability (Figure 4E,F). We found that, as expected, breathability depended strongly on porosity (correlation coefficient, r = 0.929 for woven fabrics, r = 0.894 for knit fabrics). Although the correlation between breathability and porosity was high for both woven and knit fabrics, the slopes of the regression lines were different. For the same porosity, knit fabrics had higher breathability than woven fabrics (Figure 4E). This is due to the porosity being measured under static conditions, whereas breathability measurement involved applying air pressure across the fabrics. In the latter case, fabrics can stretch due to air pressure. Knit fabrics are overall more stretchable than woven fabrics, and therefore have higher breathability for a given static porosity. Breathability was also anti-correlated with droplet blocking efficiency (r = −0.820). For example, a single layer of T-shirt (fabric 6) had high breathability (7.23 ±0.55 mm/Pa·s) and relatively low blocking efficiency (median 83.1%, minimum 42.0%). The addition of a second layer increased droplet blocking efficiency (median 98.1%, minimum 94%) while reducing breathability (to 3.87 ±0.06 mm/Pa·s).

The porosity-breathability and efficiency-breathability relationships shown in Figure 4E,F suggest that droplet blocking efficiency may also anti-correlate with porosity. Indeed, we found this to be the case (efficiency vs. porosity: r = −0.796 for woven fabrics, r = −0.822 for knit fabrics). Similar to the porosity-breathability relationship, the slopes of the regression lines in efficiency vs. porosity plots were different for woven and knit fabrics (Figure S3). For the same porosity (measured in static conditions), knit fabrics tend to have lower droplet blocking efficiency than woven fabrics. High-speed recordings show that fabric samples can deform and stretch as a result of the impact from high-velocity droplets (see Video S3). Since knit fabrics are overall more stretchable, their effective porosity may increase during impact, leading to lower droplet blocking efficiency.

### Possible Mechanisms of Droplet Blocking by Home Fabrics

The data presented so far demonstrates that most home fabrics with one layer can block both high and low impact droplets reasonably well. With 2 or 3 layers, their blocking efficiency becomes comparable to that of medical masks while still having similar or higher breathability. However, the materials of the medical mask and that of the home fabrics are very different. How do the home fabrics achieve their blocking efficiency? While porosity and stretchability play a role, as discussed above, we also observed differences between the medical mask material and home fabrics in terms of wetting and water soaking behavior. Commercially manufactured medical masks (Figure 1A,B) use 3 layers of hydrophobic fabric (non-woven plastic material, *e.g*., polypropylene) with a high contact angle (Figure 5A, Video S6). They do not wet. This hydrophobicity may be one of the factors that offer them high droplet blocking efficiency. Most home fabrics, on the other hand, are hydrophilic to different degrees. They soak water (Video S6).

**Figure 5.**
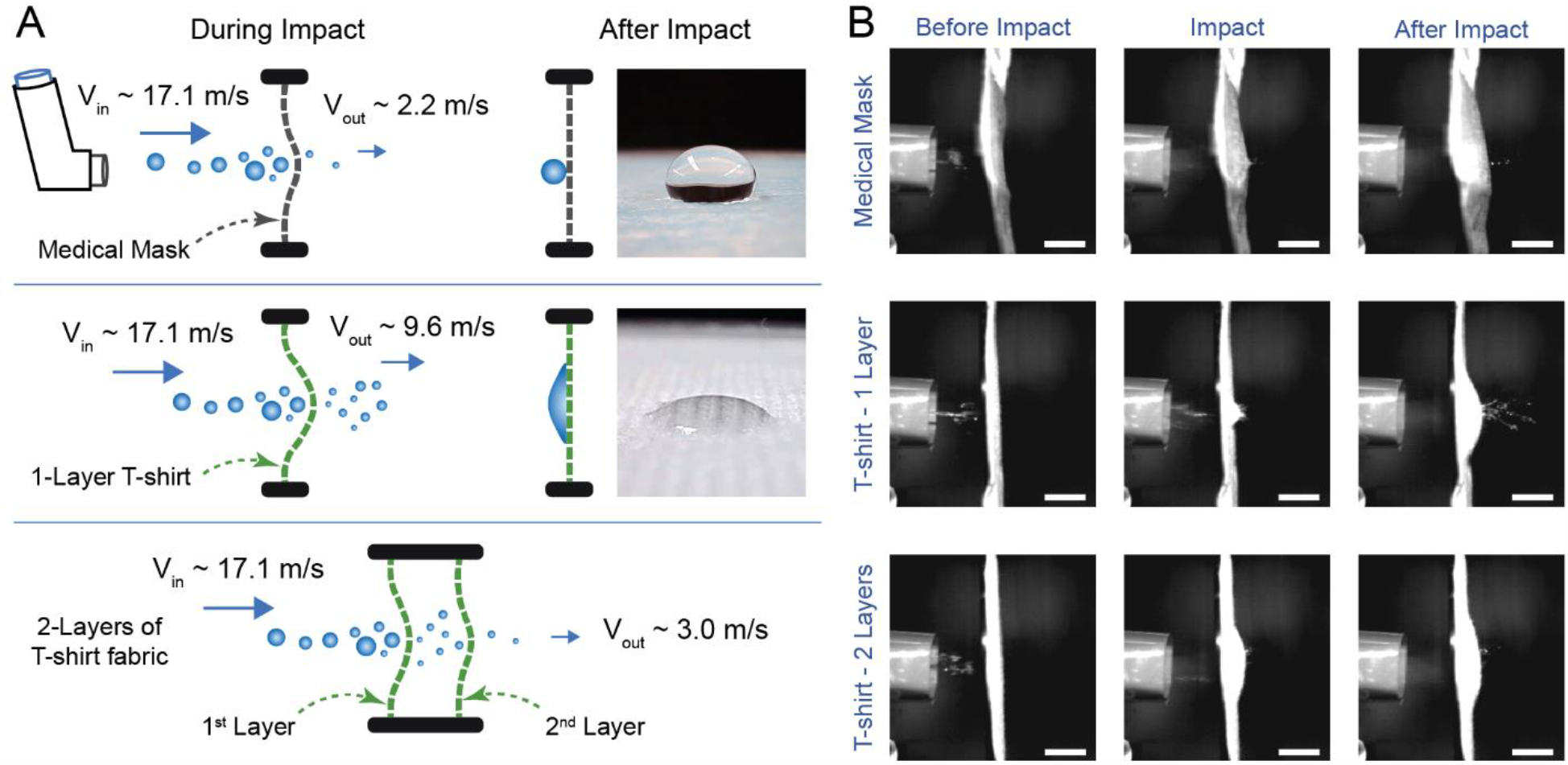
Analysis of high impact droplet resistance by T-shirt fabric and medical mask. (A) Schematic representation of the processes during high-speed impact of droplets on medical mask and cotton T-shirt fabric (fabric 6). Note that the gap between the 2-layered fabric is exaggerated to highlight the droplets between the layers. Images show water droplets on medical mask and T-shirt fabric. While the mask is highly hydrophobic, T-shirt fabric is hydrophilic. (B) Impact response of various samples. Top row: medical mask, middle row: 1 layer of T-shirt fabric, bottom row: 2 layers of T-shirt fabric. It is apparent that mask material does not bend much, compared to the T-shirt fabric samples that undergo extensive bending deformation due to impact. Scale bars: 10 mm.

To better understand the underlying mechanism of droplet blocking by hydrophilic home fabrics, we recorded high-speed videos of the incident droplets from the inhaler and subsequent transmission through the medical mask, as well as 1 and 2 layers of T-shirt (fabric 6) (Figure 5B, Videos S2-S4). In all cases, the samples were attached to a 40 mm-diameter wire ring which was placed 25 mm from the inhaler nozzle. Image analysis reveals that the droplets impact and push on the fabric (medical mask or T-shirt fabric) with a median velocity of 17.1 m/s. They also bend and stretch the fabrics. A few droplets penetrate the fabrics and split into smaller droplets as they exit. With a single layer of T-shirt fabric, a significant number of small droplets pass through (Figure 5B, Video S3), whereas the medical mask allows only a few small droplets to go through (Figure 5B, Video S2). The median velocity of the small droplets leaving the single layer of T-shirt fabric was 9.6 m/s, significantly lower than their incident velocity of 17.1 m/s (Figure 2C, p < 0.001, two-sample t-test). The kinetic energy of the incident droplets is spent deforming the fabric (bending and stretching) and in splitting the droplets (Figure 5A). The exiting small droplets do not have much momentum left to impact the second layer, if present. The second layer might also soak many of them. Indeed, high-speed imaging reveals very few droplets exiting the 2-layered T-shirt fabric sample (Figure 5B, Video S4). This reduction of momentum and the ability to soak water may explain the high blocking efficiency of 2-layered T-shirt fabric sample (median 98.1%, minimum 94.0%). Thus, energy dissipation and soaking appear to be key mechanisms that contribute to the droplet blocking efficiency of home fabrics. After completion of the impact and partial transmission, the T-shirt fabric may soak and retain the remaining droplet volume, whereas hydrophobic medical mask does not absorb any liquid. Large droplets may just roll down the medical mask by gravity.

The above scenario involving high-speed droplets applies to users coughing and sneezing on the mask. As for a mask user receiving the droplets arriving with low momentum (Video S5), the droplets are likely to be soaked by the outer layer before even reaching the inner layer, giving a high droplet blocking efficiency, similar to that of the medical mask (Figure 3B, Table 2). Thus, hydrophilicity or soaking ability of home fabrics provides an alternative mechanism of droplet blocking, in contrast to the water-repellant hydrophobic medical masks.

### Home Test of Water Permeability

The techniques we developed and used in this study to measure breathability and droplet blocking efficiency involved specialized laboratory equipment not available to the general public. We therefore sought a simple method that could be used at home to help compare different fabric choices. Here, we describe a simple test of water permeability and assess correlations between its result and breathability and droplet blocking efficiency.

We pre-wetted the fabrics with water (medical mask was not used here since it did not wet), placed them onto the mouth of a bottle and tied in place with a rubber band. The bottle contained a cup (∼250 ml) of water and had a small hole punched at mid-height to equilibrate pressure inside the bottle to atmospheric pressure. Next, we gently flipped the bottle to allow water to drain through the wetted fabric (Figure 6A). We used a stopwatch to record the time that it takes water to drain, *T*_*drain*_, stopping when water stops streaming and begins to drip. Water drains faster through fabrics with higher water permeability. Thus, the inverse of the draining time, 1/*T*_*drain*_, offers a measure of water permeability. We found that 1/*T*_*drain*_ correlates strongly with breathability, which is a measure of air permeability (r = 0.943, Figure 6B). This strong correlation implies that both measures (water and air permeability) dependent on the same fabric properties, among which porosity is most likely a dominant one. Given that breathability and droplet blocking efficiency were anti-correlated (Figure 4F), we expect that water permeability should also anti-correlate with droplet blocking efficiency. Indeed, a comparison of median droplet blocking efficiency and 1/*T*_*drain*_ revealed this to be the case (r = −0.828, Figure 6C).

**Figure 6.**
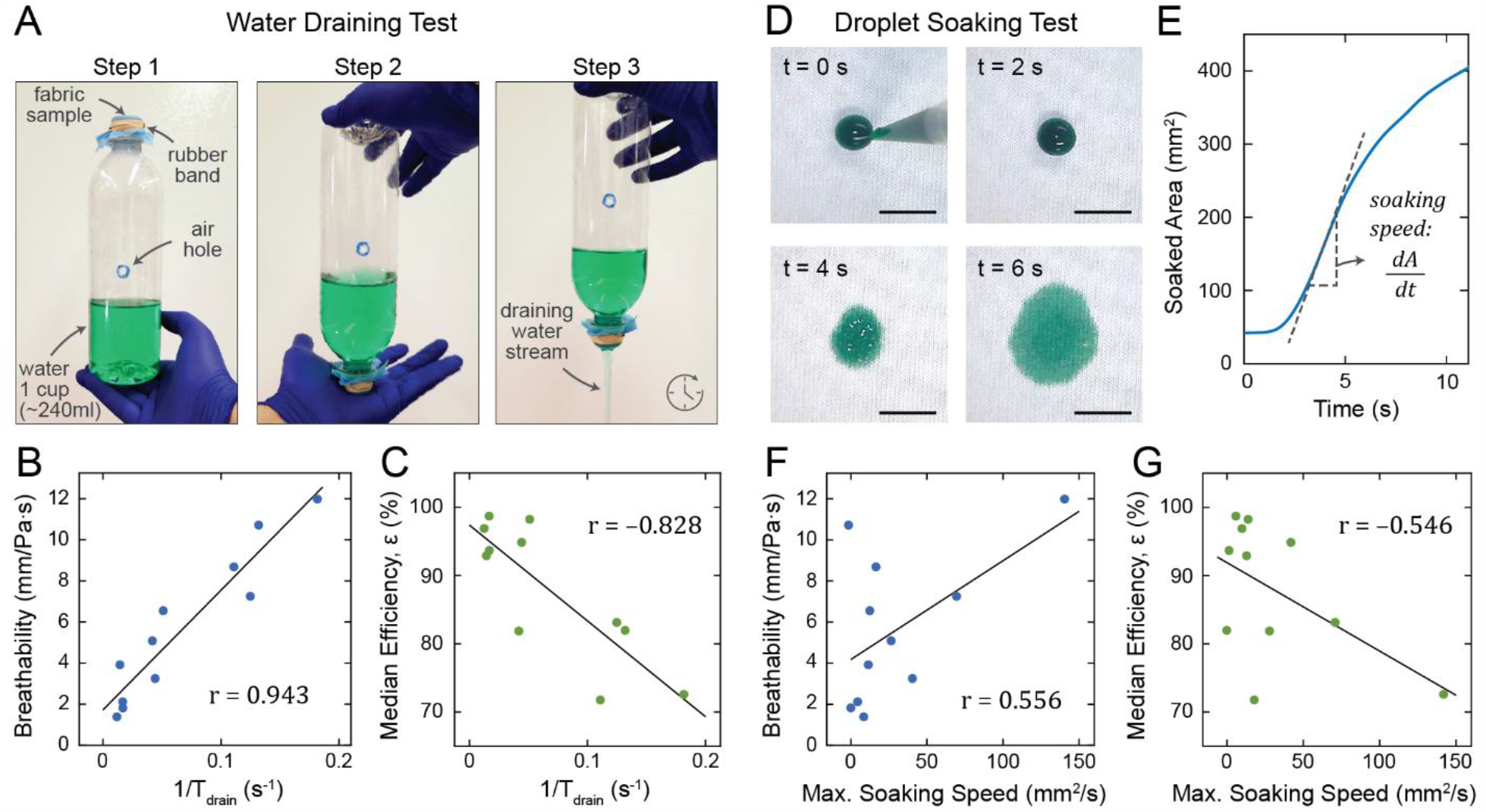
Water permeability and water soaking tests. (A) Step-by-step demonstration of the water draining test. Green food coloring is added to the water for clear visualization. Plots with regression lines of (B) breathability vs. inverse of the draining time and (C) median droplet blocking efficiency vs. inverse of the draining time. (D) Snapshots from water soaking test on T-shirt fabric (fabric 6). Green food coloring is added to provide contrast. Scale bars: 10 mm. (E) Soaked area vs. time plot for the T-shirt fabric. Soaking speed, computed as the time derivative of soaked area, corresponds to the slope. Plots with regression lines of (F) breathability vs. maximum soaking speed and (G) median droplet blocking efficiency vs. maximum soaking speed.

The simplicity and availability of this water draining test – requiring only a water bottle, rubber band, and stopwatch – provides a means by which individuals at home can assess their choices of household fabrics for masks. The result of this test, 1/*T*_*drain*_, correlates (or anti-correlates) reasonably strongly with breathability and droplet blocking efficiency that were measured in a laboratory setting (Figure 6B,C). Nonetheless, due to the approximate nature of the method, we suggest that those who choose to perform the water draining test at home to indirectly estimate breathability and droplet blocking efficiency should use it only to evaluate *relative measures* between fabrics and not to determine absolute values of breathability or efficiency.

### Water Soaking Ability of Home Fabrics

As discussed above, home fabrics and medical mask vary in terms of their water soaking behavior (Video S6). We quantified the soaking ability of each fabric and the medical mask by dispensing a small droplet (100 µl) of water on dry samples, and video recorded the soaking process to assess the relative water soaking ability of the 11 fabrics, and to explore any correlation between soaking behavior and droplet blocking efficiency. Food coloring was added to the water to provide contrast and allow us to visualize the soaked area (Figure 6D). To quantify the water soaking speed, we performed image analysis and measured the soaked fabric area as a function of time (Figure 6E, Figure S4). Water soaking speed was then computed as the time derivative of the soaked area (Figure S4). We found that the maximum soaking speed is weakly correlated with breathability (r = 0.556, Figure 6F) and weakly anti-correlated with median droplet blocking efficiency (r = −0.546, Figure 6G) for single layers of fabric. These weak correlations are not surprising. Blocking of high momentum droplets involves a short time scale (sub-millisecond) processes, whereas soaking takes longer time (seconds to minutes). Similarly, breathability measurements involved forcing dry air through the fabric, whereas soaking involves air-water-fabric interfaces and their relative energies.

Despite the weak correlations, it is conceivable that soaking ability of home fabrics may contribute to droplet blocking efficiency of home fabrics when used in multiple layers. The first layer may allow transmission of a significant fraction of the incident droplet volume, but it reduces their velocity significantly (Figure 2C). Hence, the second layer receives low momentum droplets and can employ its soaking ability to enhance blocking efficiency. This may explain the significant increase in the blocking efficiency of multi-layered fabrics compared to their single-layered counterpart.

### Remarks

Regarding the practical use of home-made face coverings, the properties of home fabrics have several important implications aside from breathability and droplet blocking efficiency. Microscale and nanoscale properties, such as microstructure and surface chemistry, of the fabrics contribute to their wetting behavior, which can be modified by engineering these properties.^35^ Home fabrics can be washed and reused, as opposed to medical masks that are disposable. Medical masks are typically made of non-biodegradable plastics such as polypropylene. Their widespread use during a pandemic can therefore pose an environmental burden. The ability to wash and reuse home-made face coverings offers an advantage in terms of reducing waste and pollution. Washing home-made face covering is also necessary for decontamination. As we have discussed above, most home fabrics are hydrophilic (water soaking). In contrast to the highly hydrophobic medical mask material, home fabrics can soak and hold the droplets. While this holding ability of hydrophilic fabrics may accrue possible benefits in terms of droplet blocking performance of home-made masks, it also means that the fabrics can retain the viruses that were in the soaked droplets. Home-made masks must therefore be washed regularly to decontaminate, either by laundry machine at warmest temperature setting, or by hand using water and household disinfectants such as bleach.^11,17,18^

The results of droplet blocking efficiency we have presented here complement the existing knowledge from studies on the aerosol filtration efficiency of household fabrics.^12–16^ The method commonly employed in these studies, as well as in the standard testing of commercial facemasks and respirators, involves challenging the sample by a pressure-driven airflow containing aerosols with particle sizes usually within the ∼10 nm to ∼10 µm range, and using a particle counter to measure and compare the number of particles upstream and downstream of the fabric.^36^ A recent study of the performance of common household fabrics against aerosols with particle sizes within 10 nm to 6 µm range reported several findings that align with ours: Fabrics with lower porosity (tighter weave) performed better than those with higher porosity. Multiple layers of fabric were highly effective (> 90% filtration efficiency in most cases).^20^ In our experiments, the fabrics were challenged with droplets with sizes of up to ∼1 mm, significantly larger than the particles or droplets used in aerosol filtration studies. As our results indicate, household fabrics can also be effective at blocking such large droplets, especially with multiple layers.

Blocking of both small aerosol particles and large droplets by fabrics can be considered within the general theoretical framework of advective-diffusive transport through a porous barrier. The performance of the barrier can depend on which mode of transport dominates. In our experiments, the motion of droplets towards the fabric samples can be described as projectile motion with relatively high velocity. Similarly, in the previous aerosol filtration studies, aerosol particles travel towards the fabric with momentum imparted upon them by pressure-driven bulk airflow. Thus, in both cases, advective transport plays a dominant role, as opposed to diffuse transport which may occur, for instance, due to osmotic pressure. In most practical situations of facemask use, advective transport is dominant. Droplets or aerosols released by a mask wearer during sneezing, coughing, or speaking will impact the mask with high velocity. In situations where a facemask is intended for blocking incoming droplets or aerosol particles, advective motion is also expected due to ambient airflow or flow generated by inhalation. We therefore expect our results, and the findings of previous aerosol filtration studies, to be applicable in these scenarios. Situations where diffusive transport dominates may arise in confined spaces with high concentration of airborne particles but with minimal ambient airflow, thereby creating osmotic pressure across the mask. Future studies are required to assess the performance of facemasks in such situations involving predominantly diffusive transport.

While fabric porosity appears to play a role in the blocking of both aerosols and larger droplets, the mechanisms of penetration may be different. Aerosols with particles in the nanometer to micrometer range may pass through the fabric pores without interacting with the fabric. Thus, lower porosity (as a fraction of the total fabric area) will result in higher filtration efficiency. With multiple layers of fabric, the net effective porosity is likely to decrease significantly due to misalignment of the pores of the separate layers. This can significantly increase filtration efficiency against small particles. Droplets, on the other hand, can squeeze and flow through the pores. Lower porosity can still be advantageous since squeezing through smaller pores imposes higher shear stress, therefore requiring more energy. For droplets squeezing through pores, the energies of air-water-fabric interfaces also play a crucial role. As we have discussed above, household fabrics can vary in their degree of hydrophilicity and water absorption properties. A more hydrophilic fabric can facilitate the flow of water through the pores, whereas squeezing through the pores of a hydrophobic fabric would require more energy. Again, a multi-layered fabric barrier will likely misalign the pores of the separate layers, decreasing the effective porosity, and significantly increasing the blocking efficiency.

Although the relative contributions of aerosols and larger droplets to the spread of COVID-19 are not fully understood, our results, taken together with those of aerosol filtration studies, suggest that home-made face coverings may have considerable efficacy in blocking both. Furthermore, a recent modeling study suggests that extensive adoption of even relatively ineffective face masks may meaningfully reduce community transmission of COVID-19 and decrease peak hospitalizations and deaths.^37^ Mask use reduces transmission rate in a nearly linear proportion to the product of mask effectiveness (as a fraction of potentially infectious contacts blocked) and coverage rate (as a fraction of the general population), while the impact on epidemiologic outcomes (death, hospitalizations) is highly nonlinear.^37^ As a result, it is anticipated that face coverings made from household fabrics can play a vital role as a mitigating strategy. Furthermore, cloth face coverings can be made more effective by ensuring proper sealing against the face contour. For example, a recent study reported that adding a layer of nylon stocking over the masks minimized the air flow around the edges of the masks and improved particle filtration efficiency for both home-made and commercial masks.^38^ As an alternative measure to ensure a better fit, an elastomeric net or half-mask can be used over the mask.^39^

## Conclusions

In this study, we asked whether face coverings made from home fabrics can be effective against the dissemination of droplets carrying 100 nm size infectious viruses, such as SARS-CoV-2, and if so, will their droplet blocking efficiency be comparable to that of a commercial medical mask. We studied a diverse set of 11 common household fabrics with varying fiber types and constructions. We quantified the ability of these fabrics to block high velocity droplets similar to those that may be released by sneezing, coughing, or speaking. We found that all of these fabrics have considerable efficiency at blocking high velocity droplets, even as a single layer. With 2 or 3 layers, even highly permeable fabrics, such as T-shirt cloth, achieve droplet blocking efficiency that is similar to that of a medical mask, while still maintaining comparable breathability.

For low-velocity droplets, we found that blocking efficiency of T-shirt fabric is much higher compared to that for high velocity droplets. A scenario involving low-velocity droplets may arise when a mask user receives droplets released by an infected individual. It thus follows that a 2 or 3-layered home-made mask with most common fabrics may help prevent the dissemination of droplets by infected individuals, and protect healthy individuals from inhaling droplets, with efficiencies similar to that of commercial medical masks.

We also measured the breathability of the home fabrics. Breathability is a critical parameter for any mask design. A high-efficiency mask material with low breathability may let air flow through the gaps between the mask and face, allowing droplets to enter or exit the respiratory system. Home-made face coverings from such low-breathability fabrics would only give a false sense of protection.

Considering our results within the context of recent literature on aerosol filtration efficiency of home fabrics and epidemiological modeling of the potential impacts of mask use, we conclude that during pandemics and mask shortages, home-made face coverings with multiple layers can be effective against transmission of respiratory infection through droplets. Mask wearing by all individuals, supported by proper education and training of mask making and appropriate usage, can be an effective strategy in conjunction with social distancing, testing and contact tracing, and other interventions to reduce disease transmission.

## Methods

### Fabric Characterization

To quantify fabric weight (mass per unit area), rectangular samples of known area were cut from each fabric and medical mask using a paper trimmer and their masses were measured using a high precision lab scale. Fabric construction (woven, knit, or napped) was determined by visual inspection. Fiber contents were noted from the cloth labels. To measure thread count, close-up views of the fabric samples (see Figure 1) were imaged using a digital camera (MS100 USB Microscope, Teslong), and the thread count was calculated as the sum of the number of threads per unit length in weft and warp directions. Thread count measurement was not applicable to the medical mask and dishcloth due to their non-woven and napped fabric construction, respectively. To measure fabric porosity, a digital image analysis method was used (see Figure S5), adapted from previous studies.^40,41^ Samples of woven and knit fabrics were imaged while being illuminated from the backside by diffused light. Light that passed through the fabric was used to identify the pores. Backside illumination intensity was kept constant and 9 samples were imaged for each fabric. Images were then analyzed in MATLAB as follows: Images were converted to greyscale. A median filter was applied to reduce noise. Filtered images were converted to binary by applying a threshold at 90% intensity; hence, pores were identified as pixels with intensity values in the top 10^th^ percentile. Binarized images were analyzed to calculate porosity as the ratio of the number of bright pixels to the total number of pixels in the image (Figure S5). For the medical mask and dishcloth, this method did not produce images with sufficient contrast; therefore porosity measurement for these fabrics was omitted.

### Droplet Generation and Characterization

We used a metered-dose inhaler (HFA-propelled, 210 sprays, GlaxoSmithKline) and loaded its nozzle with 10 µl of distilled water to generate droplets. To measure droplet velocity, we recorded videos of the ejected droplets at 10,000 frames per second (fps) using a high-speed camera (FASTCAM-ultima APX, Photron). Videos of 4 separate sprays were recorded and analyzed in ImageJ. Droplet positions were tracked manually across consecutive frames and velocity was calculated by dividing the distance travelled by the elapsed time. To estimate ejected droplet size, we used a separate imaging setup (4MP 2560×1600 CMOS camera, Phantom) which offered higher spatial resolution with a lower frame rate (400 fps). Droplets were illuminated with a laser (50 mJ Terra PIV dual cavity YLF laser, Amplitude) for clear visualization. Images of ejected droplets from 3 separate sprays were analyzed using ImageJ and MATLAB. Captured images were converted to binary and droplets were identified as objects (8-connected components) in the binary image. Droplet diameter was calculated as the ‘equivalent diameter’ of a circle with the same area as that of the imaged droplet. Using this imaging and analysis method, we were able to detect droplets with diameters greater than ∼0.1 mm (Figure S1). For measurement of droplet size at 300 mm from the inhaler nozzle (low momentum droplets), the droplets were collected on a polystyrene dish placed perpendicular to the spray direction. The dish was then imaged immediately using brightfield microscopy. ImageJ and MATLAB were used as before to convert images to binary and measure the equivalent diameter of the landed droplets. Since water contact angle on polystyrene is about 87°,^42^ we approximated the landed droplets as half-spheres, and calculated the diameters of corresponding incoming droplets using volume conservation (Figure S1).

### Droplet Challenge Tests

To challenge the fabric samples with droplets, we placed the inhaler at mid-height of an acrylic channel open at both ends. The channel prevents air flow within the room from interfering with the tests. Droplets were generated using a suspension of 100 nm-diameter red fluorescent beads (ex/em 580/605 nm, Invitrogen, catalog #F8801) diluted in distilled water. For testing the fabric samples, we first coated the bottom of a petri dish with 1 ml of warm gelatin solution (5% wt/v) which was prepared by dissolving powdered gelatin (Sigma Aldrich, catalog #G9391) in distilled water. Gelatin forms a hydrogel at and below room temperature and melts at higher temperatures.^43^ We let the gelatin solution gel inside the petri dishes at 4°C, then covered the dishes by attaching the fabric cut-outs to the rim of the dish using double-sided tape. We placed samples inside the acrylic channel, at mid-height, 25 mm or 300 mm away from the inhaler nozzle, to challenge with high or low-velocity droplets, respectively. In each test, after the inhaler was pressed and droplets containing fluorescent beads impacted the fabric samples, droplets that penetrated the fabric samples were collected on the gelatin layer. Similarly, droplets were collected on separate gelatin-covered petri dishes without any fabric barrier, as control. Next, we warmed the samples to 37°C in an incubator to liquefy the gelatin layer and allow the beads to dissolve into the gelatin solution. This mixture was transferred to a vial, vortexed for 20 s, and then sonicated for at least 30 min to uniformly disperse the fluorescent beads in solution. This homogenized bead-gelatin mixture was re-gelled at 4°C. The beads were thus frozen in place in the hydrogel.

### Fluorescent Bead Counting and Validation

We imaged the gels containing fluorescent beads on a confocal laser scanning microscope (LSM 710, Zeiss) using a 40X water-immersion lens (NA = 1.2). For each sample, we picked five random fields of view and took z-stacks with 10 µm spacing (to ensure the same set of beads were not imaged twice) and 20 to 30 slices, resulting in 100 to 150 images per sample. The bead distribution was reasonably uniform in plane and across the gel thickness. Confocal microscopy images were analyzed in MATLAB. Images were converted to binary by applying a threshold at 50% intensity. Beads were identified as objects (8-connected components) in binary images, and the average number of beads per image, 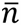, was calculated from the 100-150 images for each separate test. To validate that 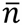 can be used as an accurate predictor of the bead density in the gelatin mixture, we prepared gelatin mixtures with known bead densities by serial dilution of the original bead solution (known bead density provided by vendor). We then gelled these solutions, performed confocal imaging, and computed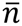, averaged over 100 images for each density tested. For samples with 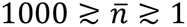, the average number of beads per image, 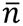, correlated very strongly with the known bead density in the mixture. Bead densities corresponding to 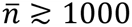 were not tested. For 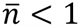, there was significant deviation from the regression model (Figure S2). Thus, for all droplet blocking efficiency measurements, we took 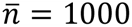 and 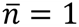 as our upper and lower detection limits, respectively.

### Breathability Measurement

Our plug flow apparatus (Figure 4B) consists of an acrylic tube with 100 mm inner diameter, a pump on one end, and a set of long aluminum tubes aligned within the acrylic tube on the other end which help stabilize the flow ^32–34^. Flow through the tube can be controlled by varying the pump speed with an analog dial. We measured the pressure, *p*, and flow velocity, *v* at the mid-length of the tube using a pressure gauge (Magnehelic) and a hot-wire anemometer (Omega), respectively. A small hole in the acrylic tube was used to insert the pressure probe and the anemometer and the hole was sealed with clay dough. An O-ring at the open end of the tube was used to seal the sample fabric, ensuring that the pumped air flows through the fabric only.

## Supporting Information

### Figures S1-S5

Characterization of droplet size; validation of fluorescent particle counting method; droplet blocking efficiency vs. porosity relationship for woven and knit fabrics; measurement of water soaking speed of fabrics; measurement of fabric porosity.

### Videos S1-S6

Droplets ejected by inhaler; droplet challenge of medical mask; droplet challenge of single layer of T-shirt fabric; droplet challenge of two layers of T-shirt fabric; incident droplets at 300 mm away from the inhaler nozzle; wetting and water absorption behavior of medical mask, slow soaking fabric, and fast soaking fabric.

## Data Availability

All relevant data will be made available upon request.

## Acknowledgements

We thank Dr. Blake Johnson of the University of Illinois at Urbana-Champaign (UIUC) for helping us with the use of the plug tube apparatus to measure breathability of the fabrics, Dr. Sivaguru Mayandi and Dr. Austin Cyphersmith of Carl R. Woese Institute for Genomic Biology, UIUC for their help with using the confocal imaging facility. Finally, we thank Dr. Faruque Ahmed of CDC for providing valuable guidance in conducting the research.

## References

(1) Nicas, M., Nazaroff, W. W., Hubbard, A. Toward Understanding the Risk of Secondary Airborne Infection: Emission of Respirable Pathogens. J. Occup. Environ. Hyg. 2005, 2 (3), 143–154.

(2) Killingley, B., Nguyen-Van-Tam, J. Routes of Influenza Transmission. Influenza and other Respiratory Viruses. Wiley-Blackwell 2013, pp 42–51.

(3) Stadnytskyi, V., Bax, C. E., Bax, A., Anfinrud, P. The Airborne Lifetime of Small Speech Droplets and Their Potential Importance in SARS-CoV-2 Transmission. Proc. Natl. Acad. Sci. 2020, 117 (22), 202006874.

(4) How Coronavirus Spreads | CDC https://www.cdc.gov/coronavirus/2019-ncov/prevent-getting-sick/how-covid-spreads.html (accessed Jun 3, 2020).

(5) Tellier, R. Review of Aerosol Transmission of Influenza A Virus. Emerging Infectious Diseases. Centers for Disease Control and Prevention (CDC) 2006, pp 1657–1662.

(6) Bourouiba, L., Dehandschoewercker, E., Bush, J. W. M. Violent Expiratory Events: On Coughing and Sneezing. J. Fluid Mech 2014, 745, 537–563.

(7) Hu, Z., Song, C., Xu, C., Jin, G., Chen, Y., Xu, X., Ma, H., Chen, W., Lin, Y., Zheng, Y., Wang, J., Hu, Z., Yi, Y., Shen, H. Clinical Characteristics of 24 Asymptomatic Infections with COVID-19 Screened among Close Contacts in Nanjing, China. Sci. China Life Sci. 2020, 1–6.

(8) Zou, L., Ruan, F., Huang, M., Liang, L., Huang, H., Hong, Z., Yu, J., Kang, M., Song, Y., Xia, J., Guo, Q., Song, T., He, J., Yen, H. L., Peiris, M., Wu, J. SARS-CoV-2 Viral Load in Upper Respiratory Specimens of Infected Patients. New England Journal of Medicine. Massachussetts Medical Society March 19, 2020, pp 1177–1179.

(9) Rothe, C., Schunk, M., Sothmann, P., Bretzel, G., Froeschl, G., Wallrauch, C., Zimmer, T., Thiel, V., Janke, C., Guggemos, W., Seilmaier, M., Drosten, C., Vollmar, P., Zwirglmaier, K., Zange, S., Wölfel, R., Hoelscher, M. Transmission of 2019-NCOV Infection from an Asymptomatic Contact in Germany. New England Journal of Medicine. Massachussetts Medical Society March 5, 2020, pp 970–971.

(10) Pan, X., Chen, D., Xia, Y., Wu, X., Li, T., Ou, X., Zhou, L., Liu, J. Asymptomatic Cases in a Family Cluster with SARS-CoV-2 Infection. The Lancet Infectious Diseases. Lancet Publishing Group April 1, 2020, pp 410–411.

(11) How to Make Cloth Face Coverings to Help Slow Spread | CDC https://www.cdc.gov/coronavirus/2019-ncov/prevent-getting-sick/how-to-make-cloth-face-covering.html (accessed Jun 3, 2020).

(12) Ahmad Chughtai, A., Seale, H., Raina MacIntyre, C. Use of Cloth Masks in the Practice of Infection Control-Evidence and Policy Gaps. Int J Infect Control 2013, 9, 3.

(13) van der Sande, M., Teunis, P., Sabel, R. Professional and Home-Made Face Masks Reduce Exposure to Respiratory Infections among the General Population. PLoS One 2008, 3 (7).

(14) Shakya, K. M., Noyes, A., Kallin, R., Peltier, R. E. Evaluating the Efficacy of Cloth Facemasks in Reducing Particulate Matter Exposure. J. Expo. Sci. Environ. Epidemiol. 2017, 27 (3), 352–357.

(15) Davies, A., Thompson, K. A., Giri, K., Kafatos, G., Walker, J., Bennett, A. Testing the Efficacy of Homemade Masks: Would They Protect in an Influenza Pandemic? Disaster Med. Public Health Prep. 2013, 7 (4), 413–418.

(16) Rengasamy, S., Eimer, B., Shaffer, R. E. Simple Respiratory Protection-Evaluation of the Filtration Performance of Cloth Masks and Common Fabric Materials Against 20-1000 Nm Size Particles. Ann. Occup. Hyg 2010, 54 (7), 789–798.

(17) Use Cloth Face Coverings to Help Slow Spread | CDC https://www.cdc.gov/coronavirus/2019-ncov/prevent-getting-sick/diy-cloth-face-coverings.html (accessed Jun 3, 2020).

(18) How to Wash a Cloth Face Covering | CDC https://www.cdc.gov/coronavirus/2019-ncov/prevent-getting-sick/how-to-wash-cloth-face-coverings.html (accessed Jun 6, 2020).

(19) Advice on the use of masks in the community, during home care and in healthcare settings in the context of the novel coronavirus (COVID-19) outbreak https://www.who.int/publications/i/item/advice-on-the-use-of-masks-in-the-community-during-home-care-and-in-healthcare-settings-in-the-context-of-the-novel-coronavirus-(2019-ncov)-outbreak (accessed Jun 6, 2020).

(20) Konda, A., Prakash, A., Moss, G. A., Schmoldt, M., Grant, G. D., Guha, S. Aerosol Filtration Efficiency of Common Fabrics Used in Respiratory Cloth Masks. ACS Nano 2020.

(21) Bourouiba, L. Turbulent Gas Clouds and Respiratory Pathogen Emissions: Potential Implications for Reducing Transmission of COVID-19. JAMA - Journal of the American Medical Association. American Medical Association 2020, pp E1–E2.

(22) Tang, J. W., Nicolle, A. D., Klettner, C. A., Pantelic, J., Wang, L., Suhaimi, A. Bin; Tan, A. Y. L., Ong, G. W. X., Su, R., Sekhar, C., Cheong, D. D. W., Tham, K. W. Airflow Dynamics of Human Jets: Sneezing and Breathing - Potential Sources of Infectious Aerosols. PLoS One 2013, 8 (4).

(23) VENTOLIN(R) HFA (albuterol sulfate) inhalation aerosol for asthma https://www.ventolin.com/ (accessed Apr 15, 2020).

(24) Xu, Z., Hickey, A. J. The Physics of Aerosol Droplet and Particle Generation from Inhalers. In Controlled Pulmonary Drug Delivery; Springer New York, 2011; pp 75–100.

(25) Dal Negro, R. W., Longo, P., Ziani, O. V., Bonadiman, L., Turco, P. Instant Velocity and Consistency of Emitted Cloud Change by the Different Levels of Canister Filling with Metered Dose Inhalers (MDIs), but Not with Soft Mist Inhalers (SMIs): A Bench Study. Multidiscip. Respir. Med. 2017, 12 (1), 1–6.

(26) BarOn, Y. M., Flamholz, A., Phillips, R., Milo, R. SARS-CoV-2 (COVID-19) by the Numbers. Elife 2020, 9.

(27) Kim, J. M., Chung, Y. S., Jo, H. J., Lee, N. J., Kim, M. S., Woo, S. H., Park, S., Kim, J. W., Kim, H. M., Han, M. G. Identification of Coronavirus Isolated from a Patient in Korea with Covid-19. Osong Public Heal. Res. Perspect. 2020, 11 (1), 3–7.

(28) Duguid, J. P. The Size and the Duration of Air-Carriage of Respiratory Droplets and Droplet-Nuclei. Epidemiol. Infect. 1946, 44 (6), 471–479.

(29) Respiratory droplets - Natural Ventilation for Infection Control in Health-Care Settings - NCBI Bookshelf https://www.ncbi.nlm.nih.gov/books/NBK143281/# (accessed Jun 3, 2020).

(30) Atkinson, J., Chartier, Y., Otaiza, F., Pessoa-Silva, C. L., Jensen, P., Li, Y., Seto, W.-H. Infection and Ventilation. In Natural Ventilation for Infection Control in Health-Care Settings., World Health Organization, 2009.

(31) Wei, J., Li, Y. Human Cough as a Two-Stage Jet and Its Role in Particle Transport. PLoS One 2017, 12 (1), e0169235.

(32) Mikhailova, N. P., Repik, E. U., Sosedko, Y. P. Optimal Control of Free-Stream Turbulence Intensity by Means of Honeycombs. Fluid Dyn. 1994, 29 (3), 429–437.

(33) Loehrke, R., Nagib, H. Experiments on Management of Free-Stream Turbulence. 1972, 113.

(34) Scheiman, J., Brookst, J. D. Comparison of Experimental and Theoretical Turbulence Reduction from Screens, Honeycomb, and Honeycomb-Screen Combinations. J. Aircr. 1981, 18 (8), 638–643.

(35) Park, S., Kim, J., Park, C. H. Superhydrophobic Textiles: Review of Theoretical Definitions, Fabrication and Functional Evaluation. J. Eng. Fiber. Fabr. 2015, 10 (4), 155892501501000401.

(36) Rengasamy, S., Shaffer, R., Williams, B., Smit, S. A Comparison of Facemask and Respirator Filtration Test Methods. J. Occup. Environ. Hyg. 2017, 14 (2), 92–103.

(37) Eikenberry, S. E., Mancuso, M., Iboi, E., Phan, T., Eikenberry, K., Kuang, Y., Kostelich, E., Gumel, A. B. To Mask or Not to Mask: Modeling the Potential for Face Mask Use by the General Public to Curtail the COVID-19 Pandemic. Infect. Dis. Model. 2020, 5, 293–308.

(38) Mueller, A. V; Eden, M. J., Oakes, J. J., Bellini, C., Fernandez, L. A. Quantitative Method for Comparative Assessment of Particle Filtration Efficiency of Fabric Masks as Alternatives to Standard Surgical Masks for PPE. medRxiv 2020.

(39) Han, D. H., Park, Y., Woo, J. J. Effect of the Tight Fitting Net on Fit Performance in Single-Use Filtering Facepieces for Koreans. Ind. Health 2018, 56 (1), 78–84.

(40) Ragab, A., Fouda, A., Deeb H E., Taleb H, A. Determination of Pore Size, Porosity and Pore Size Distribution of Woven Structures by Image Analysis Techniques. J. Text. Sci. Eng. 2017, 07 (05), 1–9.

(41) Benltoufa, S., Fayala, F., Cheikhrouhou, M., Nasrallah, S. Ben. Porosity Determination of Jersey Structure. AUTEX Research Journal 2007, 7 (1), 63–69.

(42) Li, Y., Pham, J. Q., Johnston, K. P., Green, P. F. Contact Angle of Water on Polystyrene Thin Films: Effects of CO2 Environment and Film Thickness. Langmuir 2007, 23 (19), 9785–9793.

(43) Osorio, F. A., Bilbao, E., Bustos, R., Alvarez, F. Effects of Concentration, Bloom Degree, and PH on Gelatin Melting and Gelling Temperatures Using Small Amplitude Oscillatory Rheology. Int. J. Food Prop. 2007, 10, 841–851.

